# Evaluation of Pre-operational Factors Contributing to a Dorsal Column Stimulator Explantation

**DOI:** 10.1101/2021.11.10.21266180

**Authors:** Maximilian A. Kinne, R. Scott Cowan, Alexander Knee

## Abstract

**Objectives:** Implantation of a dorsal column stimulator (DCS) for axial spine and radicular pain is a commonly performed procedure. Despite the benefits of this device to reduce pain and improve quality of life, some patients elect to have the device explanted. The purpose of this study is to describe pre-operational factors among patients who elected to have their DCS explanted and how these factors are associated with reason for explantation.

**Materials and Methods:** We conducted a retrospective descriptive study using the database of a private outpatient orthopedic clinic. We included all patients who had a DCS explanted between January 1, 2007 and June 19, 2014. Data was collected on patient demographics, past medical and back surgery history, as well as details of implantation, permanent device implantation, and subsequent explantation. Reasons for explantation were categorized as: inadequate pain control using three categories (with no device-related pain/discomfort, with device-related pain/discomfort, or inadequate pain control and patient wants MRI), or pain resolved.

**Results:** A consecutive sample of 100 subjects who underwent explantation of a DCS was identified for review. Of these 100 subjects, 14 were excluded. Based on our data, we hypothesize that sex (57% female, 43% male) degenerative disc disease (72%), previous back surgery (70%), BMI classified as overweight (subject average = 28.3), history of tobacco usage (57%), and history of narcotic use (80%) may be potential risk factors for explantation.

**Conclusions:** With respect to clinical evaluation of patients as candidates for spinal cord stimulator implantation, we cannot recommend that any of the evaluated variables be considered a contraindication to proceeding with a trial procedure. Future studies are planned to compare these data to a control group of subjects to establish risk factors predisposing individuals to explantation of a DCS.

## INTRODUCTION

Implantation of a dorsal column stimulator (DCS) for axial spine and radicular pain is a commonly performed procedure. Despite the demonstrated cost-effectiveness of DCS therapy relative to reoperation and medical management (1,2) and benefits of this device to reduce pain and improve quality of life (2), some patients elect to have the device explanted. Commonly cited reasons for explantation include pain or discomfort caused by the device (3), removing an MRI-incompatible DCS so that an MRI can be obtained (3) and lack or loss of efficacy of the device (3,4). It is not well understood why a DCS fails to control pain for these patients, as there exists no large series examining the relationship between pre-operational factors in populations that underwent an implantation of this device and a subsequent explantation. The purpose of this study is to describe pre-operational factors among patients who elected for explantation and how these factors are associated with reasons for explantation.

## MATERIALS AND METHODS

We conducted a retrospective descriptive study using the database of a private outpatient orthopedic clinic. We included all patients who had a dorsal column neurostimulator explanted between January 1, 2007 and June 19, 2014. We excluded patients whose explantation was due to infection, emergent MRI, dorsal column stimulator wound dehiscence, or worsening of comorbid symptoms from multiple sclerosis not related to axial spine and radicular pain. In terms of eligible subjects, all explantations occurred at a hospital affiliated with the study orthopedic clinic. All patients who underwent implantation of a permanent device first received a trial device implantation. Trial devices were implanted at an ambulatory surgery center, left in place for four to seven days and then explanted. If the trial device relieved greater than 50% of the patient’s pain, the patient was then offered a permanent device. The outcome of interest for explantation of a permanent device was failure of the device to control axial spine and radicular pain.

A consecutive sample of one hundred subjects who underwent explantation of a dorsal column neurostimulator was identified for chart review. Of these 100 subjects, 14 were excluded (11 due to device infection, one due to the need for an emergent MRI to rule out a brain lesion, one following dehiscence of the implantation site, and one whose multiple sclerosis worsened after implantation of the device). The remaining 86 subjects, 37 males and 49 females, underwent chart review. Subject age at the time of explantation ranged from 26-85 years.

Eligible patients were entered into a REDCap database (5) and medical records were reviewed by a premedical student trained by a senior orthopedic surgeon (RSC). The majority of records of trial and permanent device implantations performed at outside clinics were scanned into an electronic medical record; however, several records were not received or incomplete. Data was collected on patient demographics, past medical and back surgery history, as well as details of the trial device implantation, permanent device implantation, and subsequent explantation. Reasons for explantation were categorized as: inadequate pain control using three categories (with no device-related pain/discomfort, with device-related pain/discomfort, or inadequate pain control and patient wants MRI), or pain resolved.

Our analysis was descriptive in nature, and as such no hypothesis testing was conducted. Descriptive analysis included cross-tabulations of patient factors by reason for explantation. We primarily report frequencies and percentages as well as means and ranges for normally distributed continuous variables and medians and interquartile intervals (25^th^ to 75^th^ percentiles) for skewed continuous variables. Our sample size was based on all available patients at the time of study initiation. Factors that suggested at least a 10 percentage point difference in the probability of a given reason for explantation were considered to have possible clinical relevance and may be useful in generating future hypotheses.

## RESULTS

Overall, in terms of baseline characteristics, we noted that explantation was more common among females (n=49; 57%) versus males (n=37; 43%) (Table 1). In addition, a majority of patients with explantation had degenerative disk disease (n=62; 72%) or a previous back surgery (n=60; 70%). In terms of reasons for explantation, very few had their pain resolved (n=4; 5%) and most had their device explanted due to inadequate pain control (no device related pain) (n=36; 42%) or inadequate pain control (wanting an MRI) (n=35; 41%).

**Table 1:**
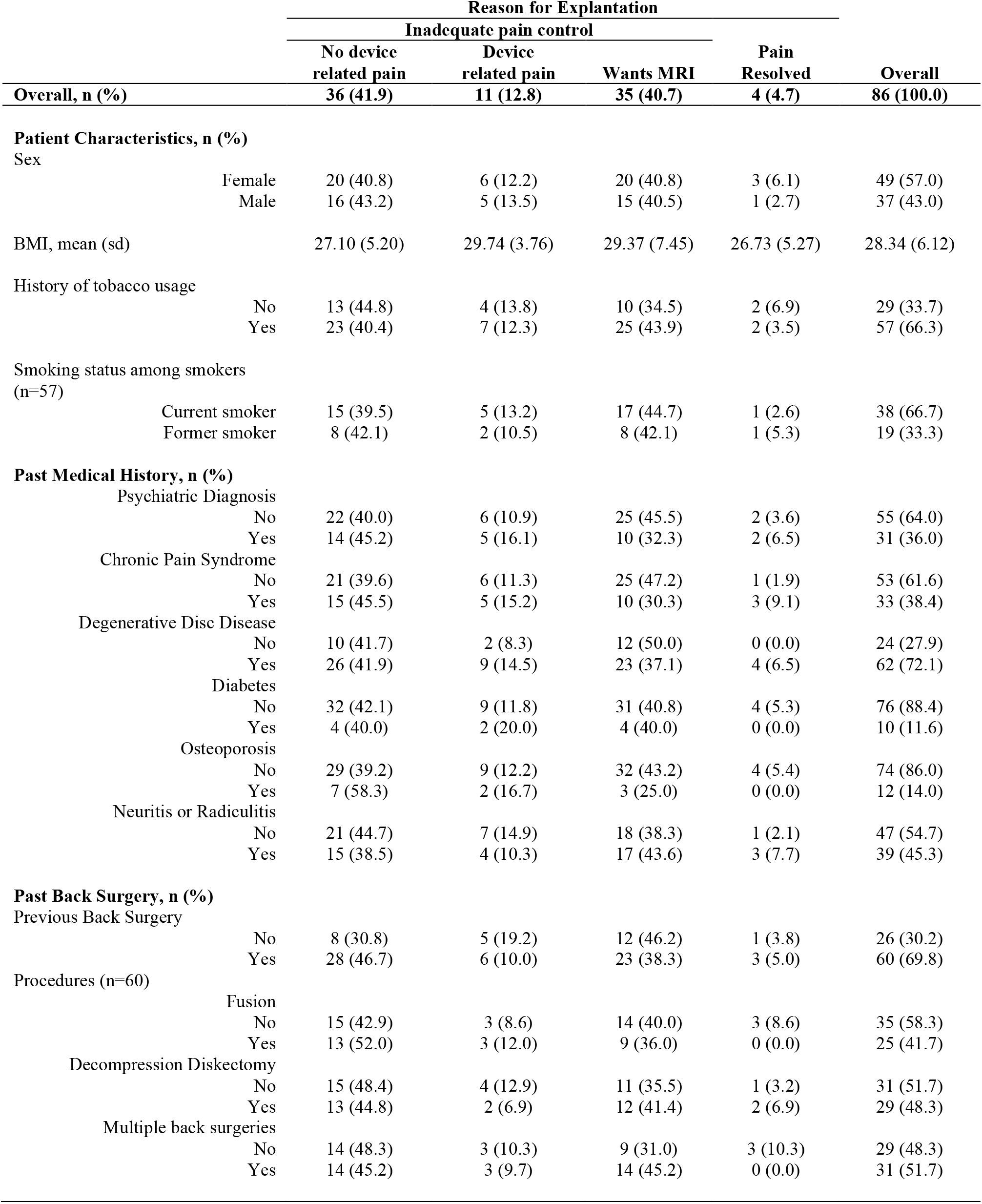
Patient Characteristics and past medical/surgical history

In terms of patient characteristics and how they predicted reasons for explantation, we did not observe many large clinically important differences as most probabilities were within 10 percentage points. In addition, due to the small sample size in the inadequate pain control (with device related pain) and pain resolved groups, comparisons were hard to evaluate. However, we noted that those with a psychiatric diagnosis were nearly 14% less likely (32% vs 46%), those with chronic pain were nearly 17% less likely (30% vs 47%) and those with degenerative disc disease were also nearly 13% less likely (37% vs 50%) to have an explantation due to inadequate pain control (wanting an MRI). Those with a prior back surgery were nearly 16% more likely to have an explantation due to inadequate pain control (no device related pain) (31% vs 47%). This difference appears to be driven by spinal fusions. However, prior back surgery was not clinically predictive for any of the other reasons for explantation.

When we evaluated characteristics of the explantation we noted that time to explantation was earliest among patients who reported inadequate pain control (with device related pain) (n=11; median days = 349) compared to approximately 600-700 days for the other groups. (Table 2). At least 75% of the patients underwent an attempt to reprogram the device, however this could be higher given that were not able to find this data in just over 20% of the records. Narcotic use from device implantation through explantation was also common (n=69; 80%), but didn’t appear to have a strong clinical association with any particular reason for explantation.

**Table 2:** Characteristics of trial & permanent device, and explantation

|  | Reason for Explantation |  |  |  |  |
| --- | --- | --- | --- | --- | --- |
|  | Inadequate pain control |  |  |  |  |
|  | No device<br>related pain | Device<br>related pain | Wants MRI | Pain Resolved | Overall |
| Overall, n (%) | 36 (41.9) | 11 (12.8) | 35 (40.7) | 4 (4.7) | 86 (100.0) |
| Trial & Permanent Device, n (%) |  |  |  |  |  |
| Trial implant device type |  |  |  |  |  |
| Medtronic | 10 (47.6) | 3 (14.3) | 8 (38.1) | 0 (0.0) | 21 (24.4) |
| St. Jude (ANS) | 16 (38.1) | 5 (11.9) | 17 (40.5) | 4 (9.5) | 42 (48.8) |
| Boston Scientific | 7 (77.8) | 1 (11.1) | 1 (11.1) | 0 (0.0) | 9 (10.5) |
| Advanced Bionics | 2 (100.0) | 0 (0.0) | 0 (0.0) | 0 (0.0) | 2 (2.3) |
| Not specified | 1 (8.3) | 2 (16.7) | 9 (75.0) | 0 (0.0) | 12 (14.0) |
| Permanent implant device type |  |  |  |  |  |
| Medtronic | 13 (46.4) | 3 (10.7) | 12 (42.9) | 0 (0.0) | 28 (32.6) |
| St. Jude (ANS) | 13 (32.5) | 6 (15.0) | 17 (42.5) | 4 (10.0) | 40 (46.5) |
| Boston Scientific | 8 (72.7) | 1 (9.1) | 2 (18.2) | 0 (0.0) | 11 (12.8) |
| Advanced Bionics | 1 (100.0) | 0 (0.0) | 0 (0.0) | 0 (0.0) | 1 (1.2) |
| Not specified | 1 (16.7) | 1 (16.7) | 4 (66.7) | 0 (0.0) | 6 (7.0) |
| Age at permanent implantation, mean (sd) | 50.5 (15.0) | 52.5 (6.4) | 47.2 (12.6) | 57.0 (15.9) | 49.8 (13.3) |
| Explantation, n (%) |  |  |  |  |  |
| Age at explantation*, mean (sd) | 52.6 (14.6) | 52.5 (10.4) | 50.5 (11.5) | 58.5 (16.7) | 52.0 (12.9) |
| Days from Permanent Implant to Explantation*, median (iqi) | 675 (375; 973) | 349 (230; 1086) | 709 (427; 1353) | 596 (342; 720) | 672 (350; 989) |
| Attempt to re-program device |  |  |  |  |  |
| Yes | 29 (45.3) | 8 (12.5) | 27 (42.2) | 0 (0.0) | 64 (74.4) |
| No | 2 (100.0) | 0 (0.0) | 0 (0.0) | 0 (0.0) | 2 (2.3) |
| Not specified | 5 (25.0) | 3 (15.0) | 8 (40.0) | 4 (20.0) | 20 (23.3) |
| Narcotic use |  |  |  |  |  |
| No | 6 (35.3) | 2 (11.8) | 8 (47.1) | 1 (5.9) | 17 (19.8) |
| Yes | 30 (43.5) | 9 (13.0) | 27 (39.1) | 3 (4.3) | 69 (80.2) |

\* Missing data: age at time of permanent implant (n=5), days from permanent implant to explantation (n=5) IQI = Interquartile interval (25th to 75th percentile)

## DISCUSSION

As there has not been any large series examining pre-operational variables in patient populations that underwent an implantation of a dorsal column neurostimulator and a subsequent explantation, the reason why a DCS fails to control pain for certain patients is not well understood. Our analysis examines pre-operational factors among patients who elected for explantation of a DCS. Collectively, 82 subjects (95%) reported inadequate pain control from a nonfunctional DCS, a larger percentage than that reported by Dupré et al (73%) (3). Based on our data, we hypothesize that sex (57% female, 43% male) may be a risk factor for explantation. Other potential risk factors include previous diagnosis of degenerative disc disease (72%), previous back surgery (70%), BMI classified as overweight (subject average = 28.3), history of tobacco usage (57%), and history of narcotic use (80%).

## CONCLUSION

Based on our study, with respect to clinical evaluation of patients as candidates for spinal cord stimulator implantation we cannot recommend that any of the evaluated variables be considered a contraindication to proceeding with a trial procedure. Future studies are planned to compare these data to a control group of subjects (patients in the same study timeframe who received an implantation of a dorsal column stimulator and did not undergo an explantation) to establish risk factors predisposing individuals to explanation of this device-important information to providers and patients alike.

## Data Availability

All data produced in the present work are contained in the manuscript.

